# Factors Influencing Smartwatch Use and Comfort with Health Data Sharing: A Sequential Mixed-Methods Study Protocol

**DOI:** 10.1101/2024.02.22.24303188

**Authors:** Anthony James Goodings, Derjung Mimi Tarn, Philip Fadahunsi, Patrick Henn, Frances Shiely, John O’Donoghue

**Affiliations:** University College Cork; University of California, Los Angeles; Imperial College London

## Abstract

**Introduction:** Smartwatches have become ubiquitous for tracking health metrics. These data sets hold substantial potential for enhancing healthcare and public health initiatives; it may be used to track chronic health conditions, detect previously undiagnosed health conditions, and better understand public health trends. By first understanding the factors influencing one’s continuous use of the device, it will be advantageous to assess factors that may influence a person’s willingness to share their individual data sets. This study seeks to comprehensively understand the factors influencing the continued use of these devices and people’s willingness to share the health data they generate.

**Methods and analysis:** A two-section online survey of smartwatch users over the age of 18 will be conducted (n≥200). The first section, based on the Expectation-Confirmation Model (ECM), will assess factors influencing continued use of smartwatches while the second section will assess willingness to share the health data generated from these devices. Survey data will be analysed descriptively and based on structural equation modelling.

Subsequently, six focus groups will be conducted to further understand the issues raised in the survey. Each focus group (n=6) will consist of 3 smartwatch users, a general practitioner, a public health specialist, and an IT specialist. Young smartwatch users (aged 18-44) will be included in three of the focus groups and middle-aged smartwatch users (aged 45-64) will be included the other three groups. This is to enhance comparison of opinions based on age groups. Data from the focus groups will be analyzed using the micro-interlocutor approach and an executive summary.

After the focus group, participants will complete a brief survey to indicate any changes in their opinions resulting from the discussion.

**Ethics and dissemination:** The results of this study will be disseminated through publication in a peer-reviewed journal, and all associated data will be deposited in a relevant, publicly accessible data repository to ensure transparency and facilitate future research endeavors.

This study was approved by the Social Research Ethic Committee (SREC), University College Cork – SREC/SOM/21062023/2.

**Strengths and limitations of this study:**

⇒ Understanding factors influencing smartwatch use and willingness to share data may provide insights on how to promote a shift from non-continuous to continuous smartwatches uses and how to overcome barriers to sharing of useful health data from smartwatches.
⇒ Examining the privacy concerns of patients regarding the sharing of health data from a smartwatch may provide additional insight into what measures are needed before integration of smartwatch data into healthcare can be normalised.
⇒ The survey will be based on the Expectation Confirmation Model which has been applied previously in similar studies and the combination of quantitative and qualitative methods will enhance the triangulation of findings.
⇒ The findings from this study will be limited to reflecting the view of individuals in Ireland and may not be directly relatable to countries with different systems of healthcare.
⇒ The discussions regarding privacy will be limited to discussing comfort with the collection of data that can currently be acquired from a smartwatch and will not be able to take into account concerns with more advanced data that these devices may be able to acquire in the future.

## Introduction

Smartwatches are becoming increasingly common, about 1-in-5 Americans have already used a wearable device in 2020[1]. It is well known that smartwatches such as the Apple Watch Series 4, 5, and 6 can monitor a wide range of vital signs such as heart rate, oxygen saturation, and simulate an accurate 3-lead electrocardiogram [2]. It has been suggested that the Apple Watch’s technology could lead to the earlier diagnosis of acute coronary disease [3]. It is considered that the Series 6 Apple Watch is a reliable way to obtain one’s oxygen saturation (SaO2) which is of great use in long term monitoring of respiratory disease [4]. Additional advances are being developed such as measuring blood-glucose using an optical sensor [5].There is a large body of research showing that the data collected by smartwatches are accurate and useful [2][3][4]. The benefits of these data sets for one’s health is reliant on the interpretation of it by the end user and/or a medical professional. Thus, these data sets hold substantial potential for purposes beyond the individual, such as public health, research, monitoring drugs for effectiveness, etc.

Wearable devices in general are made to collect data and display it to the user either on the device itself or through a companion application on a mobile device or website. In this study, it is of interest to determine how comfortable people would be sharing health data via different technological means, e.g. direct Bluetooth connection, over an internet-based service, and at different levels, e.g. with their own doctor, with the government, with independent bodies. There are concerns about the privacy and security of health data collected by wearable devices[6][7]. Despite this, a previous study found that around 57% of adults over 50 who use wearable devices would be willing to share data with researchers [8]. This highlights the need for further research to determine the factors that influence people’s comfort level with sharing health data collected by wearable devices.

### Continued Use of Smartwatches

The data that can be collected from smartwatches are only useful if people wear the devices consistently, as in the case of chronic diseases. A previous study set out a framework to evaluate continuous intention in smartwatches using the expectation-confirmation model (ECM) [9]. It identifies a variety of factors such as confirmation, perceived usefulness, satisfaction, habit, perceived usability, perceived enjoyment that influence continuance intention to use the device as shown in *Figure 1* [9]. As represented in the ECM diagram [9][10], these factors are important in the continuous use of the device and consequently the usefulness of the data collected for the purposes of medical care and public health. The specific relationships between these components are illustrated on the diagram and each component is examinable in the form of 26 targeted survey questions followed by structural equation modelling.

**Figure 1:**
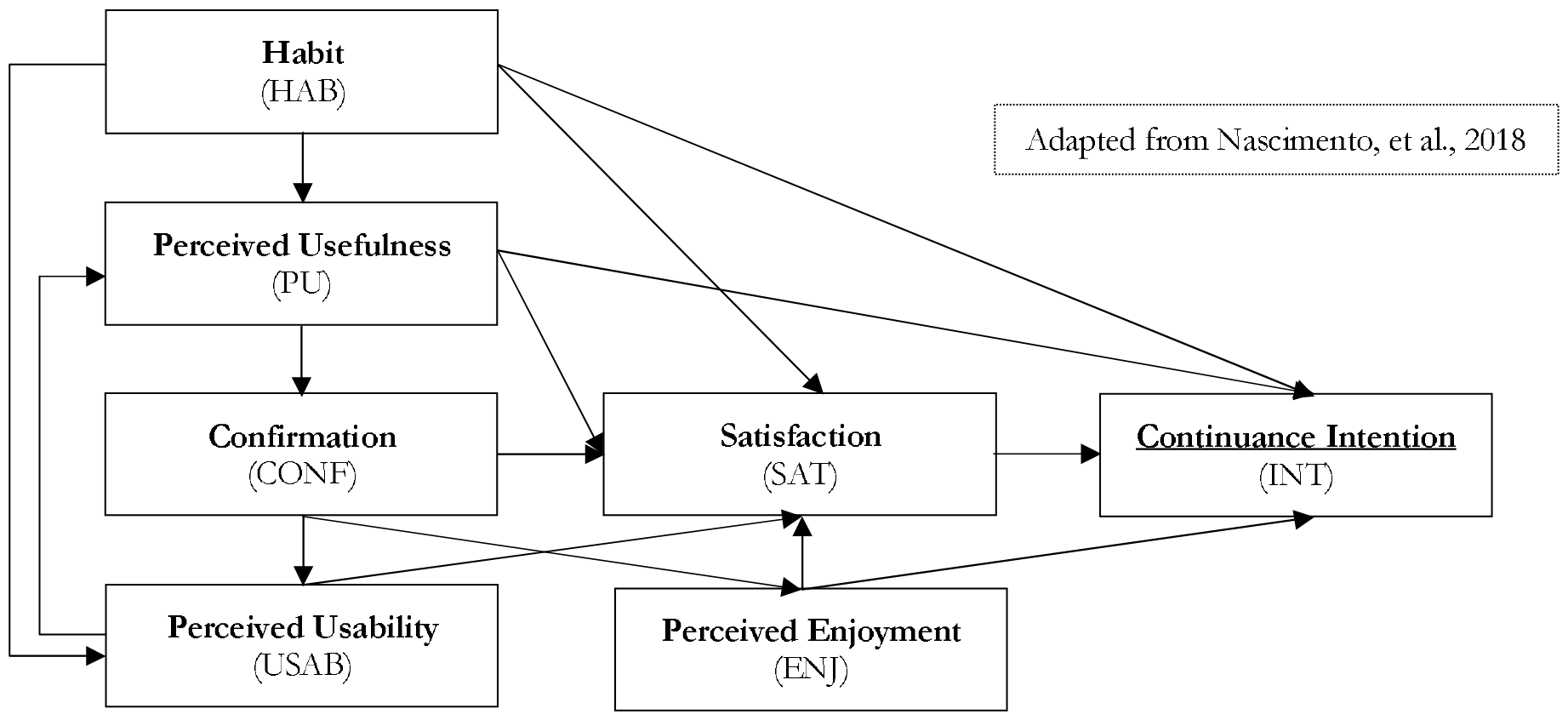
Expectation Confirmation Model

In a study about continued use of wearable activity tracker devices in older adults, it was clear that the main drivers for use were monitoring activity as a habit [11]. A key point here being that they are doing it because they enjoy it, and it brings value directly to them. It is seen as a personal device and not a medical device. Another study that focused on habit formation in smartwatch users focused on users who had used a smartwatch for at least six months, a timeframe they estimate results in the users engaging with the device without the need to think about it very much [12].

### Comfort With Smartwatch Data Sharing

Despite the low adoption of using wearable technology in primary care, there have been a number of surveys which have questioned people on whether they would like to share data with healthcare professionals. A 2018 study in Canada, found that about one-third of people share their health date with someone, and of that group one-third has shared that data with a primary care physician [13]. A survey of 2,025 American adults found that 78% of respondents who use a wearable device would choose a doctor who uses data from their wearable device [14]. A Canadian survey carried out in 2016 found that 85% of participants would share data from their wearable if their doctor requested it [15]. These figures are very encouraging when it comes to the future place of wearables in primary care. Sharing health data collected from smart devices with medical professionals is an area that warrants further research. This study aims to further the current understanding of patients’ attitudes towards sharing their data collected from smartwatches.

The surge in the popularity of smartwatches is in part for their ability to track and monitor various health metrics. However, the question of who gets access to these data sets remains a significant concern. Some users may be willing to share their smartwatch data with healthcare professionals in order to receive better care and improve their health outcomes. Many of these individuals may be hesitant to share the same data with private companies for fear of privacy violations. The role of compensation in the sharing of smartwatch data with healthcare professionals as opposed to private companies is a topic that requires further investigation. Previous research has shown that a multitude of factors influence users’ willingness to share data. One main reason is privacy concerns [16], with an increasing awareness and public interest in data privacy, any data transfer which occurs via the internet, which would be necessary when collecting data on a large scale, risks a data breach. This is a significant concern to many people, in particular those who are more familiar with technology – in general, younger adults, which make up a significant proportion of smartwatch users. Another key reason is a mistrust in researchers and institutions [17]. Generally, large corporations often lack the trust of consumers. Trust is essential in order for researchers and analysts from these companies to be given access to data by consumers. Knowledge of what the data would be used for was also identified as a key factor [18], without which many individuals would not be willing to share data. A study found that 90% of people would want to know what their data would be used for before sharing it [19].

In addition, financial incentivization could play a role in data sharing, but is has not been fully explored in the current literature. While there is little research on this exact type of data sharing, there is research on comparable topics. For instance, data sharing with e-merchants, the major consumer concern of privacy is a factor. Notably, an experimental study that tested the role of financial incentives in data sharing found that they do increase data sharing in terms of the actions taken by the participants, however, several participants did not claim to be any more interested in sharing based on the receipt of such incentives [20]. This shows that not all people are actively aware of the factors influencing their own decision to share data. This highlights the need for further research to determine the factors that influence people’s comfort level with sharing health data collected by wearable devices. This study aims to determine how comfortable people would be sharing health data via different technological means, e.g. direct Bluetooth connection, over the internet, and at different levels, e.g. with their own doctor, with the government, with independent bodies.

### Aims and Objectives

1. This study aims at better understanding of the factors influencing continued use of wearable devices.
2. This study aims to determine how comfortable people would be sharing health data via different technological means

### Research Questions

1. What factors are most important in encouraging continuous use of a wearable device?
2. What factors affect individuals’ willingness and comfort to share data with different bodies (doctor, government, private companies)?

## Methods and Analysis

### Study Design

This is a sequential mixed-methods study with both a quantitative component (a survey, appendix A), and a qualitative component (focus group, Appendix B). The organisation of the data collection process in a mixed-methods approach will allow for both the exploratory and confirmatory aspects of the study to be addressed in addition to allowing stakeholder input to be considered in focus groups [21]. Data will be collected at three points in the study: the pre-survey, six focus groups, and the post-survey for focus group participants as shown in *Figure 2*.

**Figure 2:**
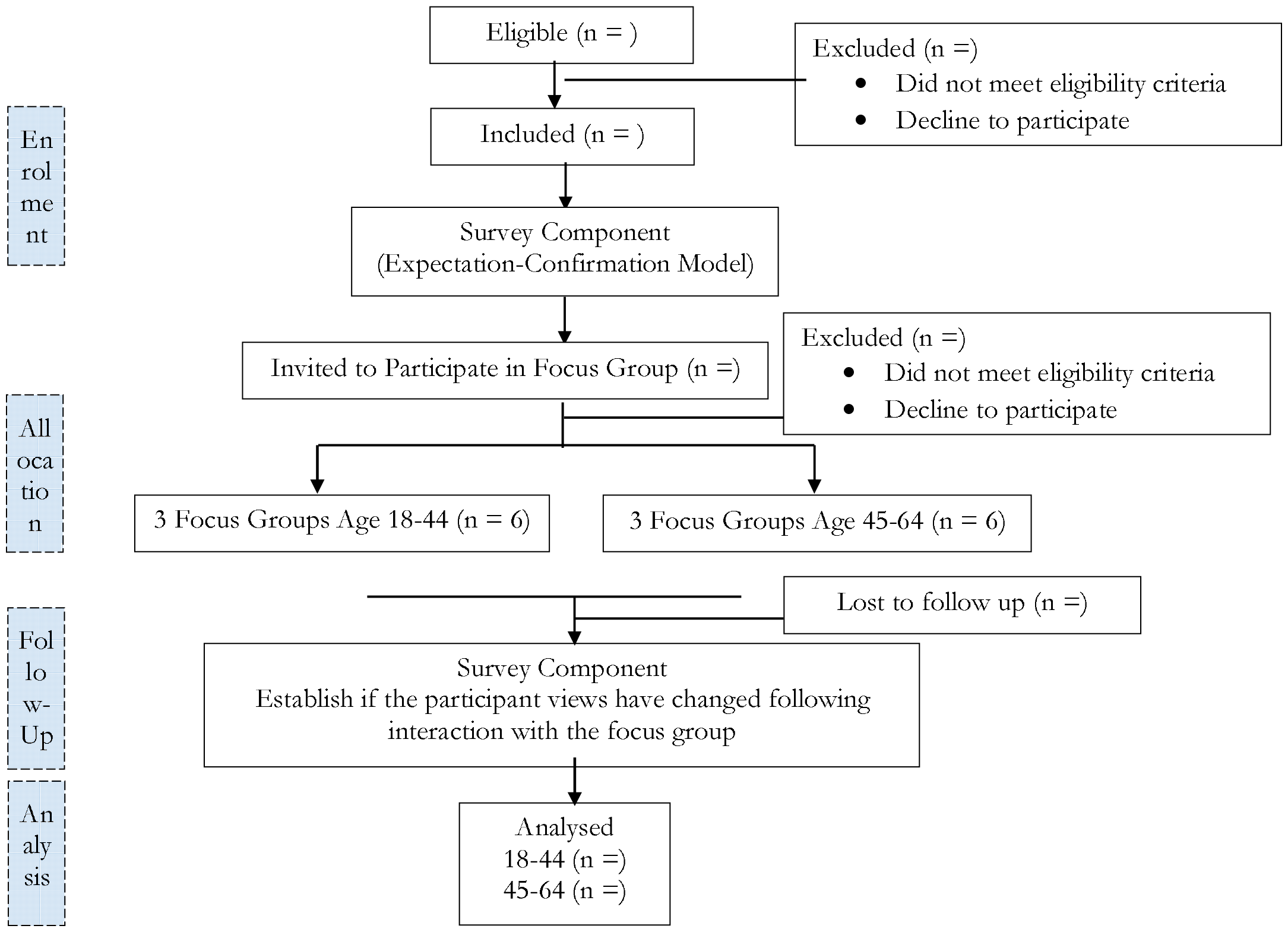
Study Procedure

A survey will be used to gain insight into the use and experiences of respondents with smartwatches as well as their willingness and comfort sharing data collected with physicians and other groups. In addition, the factors influencing the continued use of the device will be explored. The initial survey is a means of gaining insight into the participants’ experience and perceptions in the context of the *Expectation-Confirmation Model*.

Subsequently, six focus groups will be organised in which individuals of two different age groups will be asked clarifying questions based on the data collected in the survey. The participants of the focus group will be stakeholders including wearable device users, general practitioners, public health specialists (e.g. a consultant in public health medicine – a physician who has undertaken specialty training in public health, or a university instructor in the field of public health), and IT specialists as they each have a different role in the collection, storage, and use of health data collected from wearable devices. As different stakeholders bring different viewpoints, it is of significant benefit to include them in the focus groups[22] as their opinions are generally not considered when a participant is answering the survey independently. The focus groups will allow for anything that was unclear in the results of the survey to be examined further as well as get the participants to take into consideration the opinions that others had on the same topics.

Following the focus groups the participants will be given a short post-survey that will allow them to express if any of their opinions have changed due to the focus group.

### Recruitment

Smartwatch users will be recruited via convenience sampling using University College Cork survey distribution, social media (e.g. LinkedIn, local Reddit groups) and University College Cork campus radio. Participants with an interest in the research will be able to share the social media posts.

### Sample Size

The survey will aim for a minimum of 200 participants which is the recommended minimum sample for a meaningful analysis to minimize the impact of random error in structural equation modelling [23].

The main criteria for this are: **current or previous use of smartwatch** and be **over 18**. For the focus groups, the aim is to have 3 wearable device users, 1 GP, 1 IT specialist, and 1 public health expert. The number of participants was selected to be six to prevent the disintegration of the focus group into smaller groups, each holding independent discussions, that may occur when the group size is too large, typically over 12 participants [24]. A sufficient number of participants for a focus group is generally accepted to be between 6 and 8 [25]. The focus groups will be aimed at two different age groups of **smartwatch users**, one being **18-44 years old**, the other being **45-64 years old**, representing young and middle-age respectively. The age criteria will not apply to the GP, IT specialist, or the public health specialist. Investigating two different age groups separately can help determine if there is age-related difference of opinion regarding the discussed topics, which will reinforce any such pattern that may be observed from the survey data.

### Inclusion and Exclusion Criteria

The inclusion criteria for the survey include:

1. **current or previous use of smartwatch**; and,
2. be **over the age of 18 years**.

The inclusion criteria for the focussed group include:

1. Participant must be either between the **ages of 18 and 64**.
2. People with communication difficulties will not be included.

### Data Collection

The survey will be conducted online using Qualtrics. The survey (Appendix A) has two main sections. The first section, based on the expectation-confirmation model, assesses the factors influencing the continued use of smartwatches while the second section assesses the willingness of the smartwatch users to share health data with a physician and other interested parties. Following completion of the survey, data will be stored on the UCC OneDrive for business in a password protected file.

The focus groups will take place in person, with a duration of 60 minutes, session will be audio recorded and stored on a password protected device until fully transcribed. Following transcription, the audio file will be destroyed, and the transcript will be stored on the UCC OneDrive for business in a password protected file. Photos of a whiteboard draw up during the focus group will be stored on a password protected device until uploaded to the UCC OneDrive for business.

### Analysis

The survey data will be uploaded to SPSS version 29 (IBM Statistics, Armonk, New York, USA) for descriptive statistical analysis including frequency distribution, Spearman’s correlation coefficients and Cronbach’s alpha scores. The SPSS file will then be exported to SPSS Amos (IBM Statistics, Armonk, New York, USA) for structural equation modelling to test and estimate the causal relationships through the use of a mix of statistical data and qualitative causal assumptions as recommended for the adopted ECM [9].

Focus group data will be analysed using the micro-interlocutor analysis approach to gage consensus in the group. This method of focus group analysis allows for the unit of analysis to be individual as opposed to group based, it highlights contribution or lack thereof of individual members to allow consensus to be determined while concurrently reflecting individual disagreements and opinions [26]. The six focus groups will be compared with two different analysis tables modeled in the micro-interlocuter analysis framework. A short executive summary will also be provided. To maintain rigor of the qualitative data, two coders will independently analyze the data, this will help ensure objectivity and diversity of interpretation. To validate identified themes, they will be discussed with a broader group of experts to enhance the reliability of the findings.

### Data Protection

Any confidential data that is collected will be kept for 10 years on a password-protected computer and will only be accessible to those working on the study. Transcription of audio recordings will be anonymised. The original recording will be destroyed immediately following transcription. No personal identifiable data will be included in the transcribed data or subsequent publications.

### Patient and Public Involvement

The public has not yet been involved in this research. We are seeking to understand public opinion on the factors influencing smartwatch use and comfort with data sharing. We intend to first involve the public in the dissemination of the survey as well as the results.

## Ethics and dissemination

### Ethics and dissemination

The results of this study will be disseminated through publication in a peer-reviewed journal, and all associated data will be deposited in a relevant, publicly accessible data repository to ensure transparency and facilitate future research endeavors.

## Data Availability

As this is a protocol, no data is available.

## Authors’ contributions

AJG – conceptual design of the study, drafting the protocol.

FS, PH, KPF, DT – feedback and editing of the protocol.

JO’D – conceptual design of the study, feedback and editing of the protocol.

